# Lessons from applied large-scale pooling of 133,816 SARS-CoV-2 RT-PCR tests

**DOI:** 10.1101/2020.10.16.20213405

**Authors:** Netta Barak, Roni Ben-Ami, Tal Sido, Amir Perri, Aviad Shtoyer, Mila Rivkin, Tamar Licht, Ayelet Peretz, Judith Magenheim, Irit Fogel, Ayala Livneh, Yutti Daitch, Esther Oiknine-Djian, Gil Benedek, Yuval Dor, Dana G. Wolf, Moran Yassour, The Hebrew University-Hadassah COVID-19 diagnosis team

**Author notes:** Correspondence should be addressed to (D.G.W), (Y.D) and (M.Y.).

## Abstract

Pooling multiple swab samples prior to RNA extraction and RT-PCR analysis was proposed as a strategy to reduce costs and increase throughput of SARS-CoV-2 tests. However, reports on practical large-scale group testing for SARS-CoV-2 have been scant. Key open questions concern reduced sensitivity due to sample dilution; the rate of false positives; the actual efficiency (number of tests saved by pooling) and the impact of infection rate in the population on assay performance. Here we report analysis of 133,816 samples collected at April-September 2020, tested by pooling for the presence of SARS-CoV-2. We spared 76% of RNA extraction and RT-PCR tests, despite the reality of frequently changing prevalence rate (0.5%-6%). Surprisingly, we observed pooling efficiency and sensitivity that exceed theoretical predictions, which resulted from non-random distribution of positive samples in pools. Overall, the findings strongly support the use of pooling for efficient large high throughput SARS-CoV-2 testing.

## Introduction

The ongoing COVID-19 pandemic, caused by SARS-CoV-2, has resulted in substantial clinical morbidities and mortality, urging comprehensive virological testing. Major diagnostic challenges have emerged, mainly, the need for high throughput SARS-CoV-2 RT-PCR tests, aimed to detect not only symptomatic but also asymptomatic infectious viral carriers and to screen special or at-risk populations (such as health care personnel or nursing home tenants), in order to contain viral spread and guide control measures.

These diagnostic challenges together with the consequent shortage in laboratory equipment, reagents and resources call for the development of a more efficient testing strategy. One promising solution is the application of sample pooling or group testing, a well-developed field in mathematics that allows the identification of carriers in a population of N using a number of tests that is smaller than N. Group testing can alleviate the supply-chain blocks and cut costs while increasing testing throughput. Sample pooling techniques differ in the number and size of pools into which each sample is assigned. In Dorfman pooling*(1)*, which is the simplest pooling scheme, each sample is assigned to a single pool, the pools contain equal numbers of samples and samples are retested individually only if the pool’s test result is positive. In other pooling methods, samples are assigned to multiple overlapping pools in order to eliminate or at least reduce the number of retested samples*(2–5)*.

The commonly used diagnostic test for SARS-CoV-2 is based on detection of viral RNA in nasopharyngeal samples by RT-PCR amplification following RNA extraction. Pooling of samples in this context could potentially be employed at any stage along the diagnostic workflow, from pooled sample collection, to pooled RNA extraction and RT-PCR, or pooled final RT-PCR only*(2, 6–14)*, with each approach having pros and cons with regard to test saving versus logistics/delays associated with patient and sample re-testing.

We and others, have recently described the validation and early implementation of sample pooling for SARS-CoV-2 detection*(2, 6–13, 15–17)*, and, perhaps reflecting an increased confidence in this approach, the Food and Drug Administration (FDA) has already issued the first Emergency Use Authorization (EUA) for pooled testing of SARS-CoV-2 in July*(18)*. The majority of these studies have employed Dorfman pooling (with 4-32 samples per pool), and, while largely differing in protocols and stages of pooling used, have suggested sufficient diagnostic accuracy despite an expected loss of sensitivity.

When considering any of the SARS-CoV-2 pooling schemes, there are three crucial concerns: **Efficiency** - how many tests are spared in practice and its relation to the prevalence rate? **Sensitivity** - can we detect samples with lower viral load of clinical significance despite sample dilution? And **Operational feasibility** - can we technically and logistically implement the pooling scheme and quickly adapt it to changes in infection prevalence rates?

These concerns cannot be addressed by all currently reported studies, as they were conducted as a proof of concept, consisting of only hundreds to a few thousands of tested samples, examined over a short time period with a relatively constant positive samples rate (usually <1%).

Here, we describe lessons learned from a **five-month period, testing 133,816 samples using 17,945 pools**. Based on early evidence, theoretical considerations and practical limitations we chose to implement adaptive Dorfman pooling with pool sizes of 5 and 8. We evaluate the theoretical and empirical efficiency and sensitivity of our pooling approach, and its adaptation to fluctuating rates of positive samples. **Overall, we spared 76% of the PCR reactions compared with individual testing, with an acceptable reduction in sensitivity**. To our knowledge, this is the most extensive analysis, addressing key considerations of efficiency, sensitivity and feasibility in the actual reality of routine, large-scale implementation of sample pooling for SARS-CoV-2 detection.

## Results

Between mid-April and mid-September of 2020, we tested 133,816 samples in pools. One challenge to the pooling scheme stemmed from the fluctuating rates of infection during the pandemic. The infection prevalence rate of pooled samples changed considerably (despite the fact that the vast majority were obtained from asymptomatic individuals; **Figure 1A**), mandating a dynamic adaptation of the pooling scheme. In principle, at low prevalence, using fewer pools of larger pool sizes would lead to a gain in efficiency, since the majority of pools test negative. However, as prevalence increases, using a larger number of smaller-size pools would be more efficient, as every positive individual lead to retesting a smaller amount of samples (**Supplementary figure 1A**). Thus, when the prevalence rate in pooled samples increased (from ∼1% to ∼6%), we switched from 8-sample pooling to 5-sample pooling, and employed a dynamic approach thereafter (alternating the pool size between 8 and 5) to maintain optimal pooling efficiency (**Methods, Supplemental figure 1AB**).

**Figure 1.**
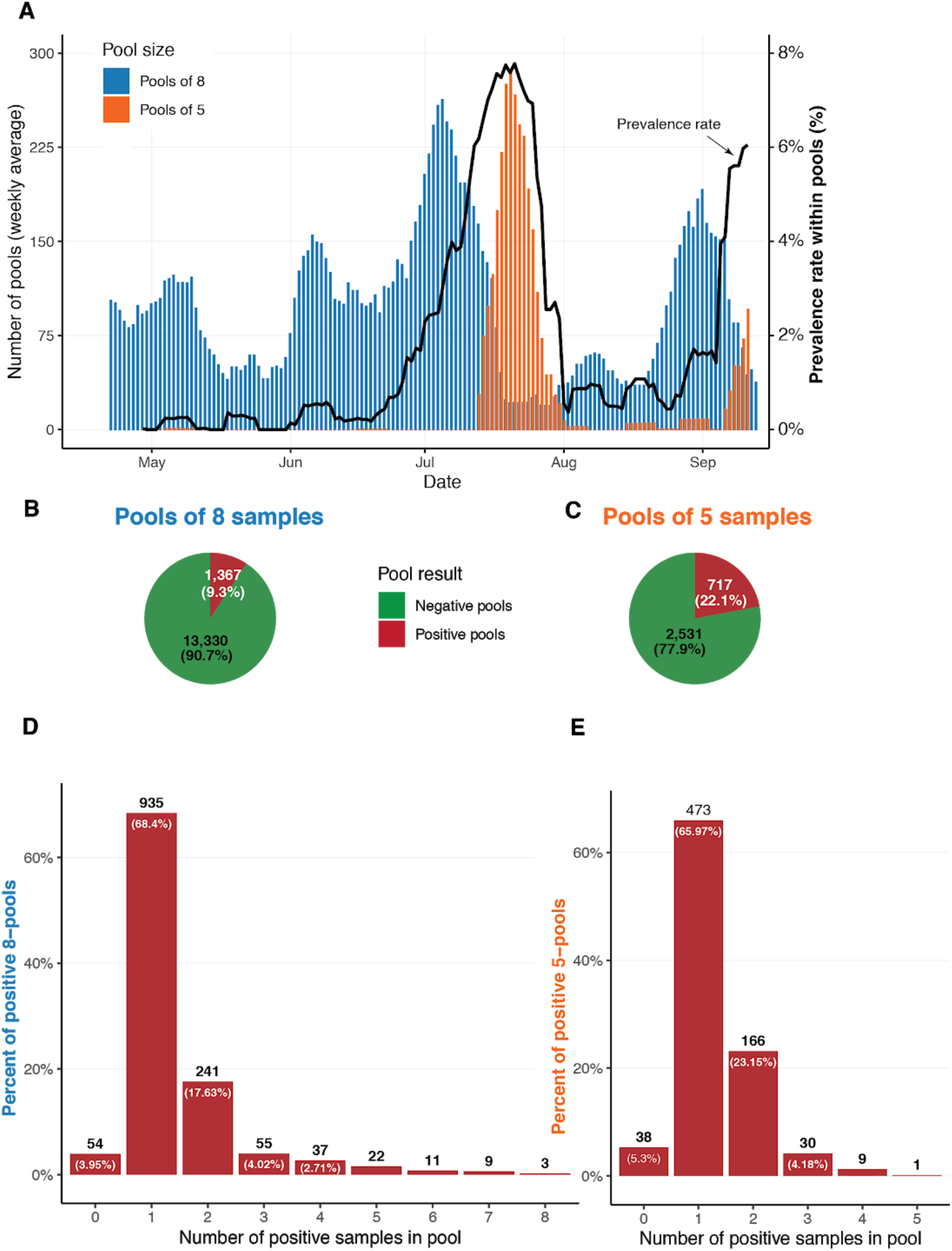
**(A)** Weekly average of 8-samples (blue) and 5-samples (orange) pools counts, together with the weekly average of the prevalence rate among pooled samples (black). **(B,C)** Pool results for 8-sample (B) and 5-sample (C) pools respectively. **(D,E)** Counts of positive pools aggregated by the number of positive samples identified within the pool, for 8-sample (D) and 5-sample (E) pools.

In total, we tested 14,697 8- and 3,248 5-sample pools, where 9.35% and 22.1% of the pools tested positive, respectively (**Figure 1B,C**). As all samples in the positive pools were re-tested individually, we could evaluate the distribution of positive samples within positive pools. While the majority (66%-68%) of the positive pools contained only one positive sample, 28%-29% of the positive pools contained two or more positive samples (**Figure 1D,E**). A small number of positive pools (3.9%-5.3%) did not yield any positive samples when their samples were re-tested individually. The viral Ct values of these pools was usually higher - with median Ct of 36.8 and 34.2 for 8- and 5-samples positive pools (respectively), while all other positive pools had median Ct of 26.9 and 26.5, respectively. This low false-positive rate (3.9%-5.3%) reflects our permissive threshold and the extra-caution taken in order to maintain the sensitivity of pooled-sample testing.

A dominant consideration in planning and evaluating the pooling approach is the efficiency, defined as the expected number of samples tested using a single RT-PCR reaction. In theory, efficiency is mostly impacted by the pool size and the prevalence rate (see **Methods, Supp Figure 1**). We calculated our *empirical efficiency* (defined as the total number of tested samples divided by the total number of actual RT-PCR reactions performed), and found it to be 4.587 and 2.377 for the 8- and 5-sample pools, respectively. Strikingly, these values are better than the expected optimal efficiency values for both the 8- and the 5-pool sizes, under the observed prevalence rates of 1.7% and 5.7%, respectively (**Table 1**). As discussed below, the reason for this supra-optimal efficiency is the non-random distribution of positive samples among pools in our real-life setting.

**Table 1:**

|  | Pools of 8 | Pools of 5 | All together |
| --- | --- | --- | --- |
| **Total number of pools** | **14,697** | **3,248** | **17,945** |
| **Number of positive pools** | **1,367** | **717** | **2084** |
| **Total number of samples** | **117,576** | **16,240** | **133,816** |
| **Number of positive samples** | **1,993** | **936** | **2,929** |
| **Total number of PCR reactions** | **25,633** | **6,833** | **32,466** |
| **Prevalence rate** | **1.7%** | **5.7%** | **2.19%** |
| **Optimal (predicted) Dorfman efficiency** | **3.9553** | **2.1891** | **NA** |
| **Our empirical efficiency** | **4.587** | **2.377** | **4.121** |

As the prevalence of infection changes, so does the pooling efficiency. Indeed, we observed fluctuations in efficiency values over time, when the empirical efficiency was higher or lower than the theoretical efficiency **(Supp Figure 1B)**. Nevertheless, across time and pool sizes we performed better than expected. Overall, we tested 133,816 samples using 32,466 RT-PCR tests with a global efficiency of 4.121, saving 101,350 (76%) reactions.

A major concern regarding sample pooling is the expected loss of sensitivity upon sample dilution. We evaluated the sensitivity in our large-scale 8-sample pooling approach, comparing the Ct value of each positive pool with the Ct value of the individually-tested positive samples within the pool. Theoretically, an 8-sample pool with a single positive sample should contain only ⅛ of the viral load, which requires 3 additional PCR cycles (log2 of the dilution factor) for detection (**Methods**). Since our PCR assay has a practical limit of sensitivity at 40 cycles, we expect pooling tests to be able to detect samples with viral Cts up to 37. Individual samples with Ct>37 are expected to be inherent false-negatives of the method. To empirically examine the theoretical loss of 3 Ct in sensitivity, we compared the pool Ct with the individual-sample Ct for 902 pools that contained only a single amplified sample (**Figure 2A**). A linear regression analysis of these data revealed a 2.9 Ct increase for the pool (R^2^=0.66, constraining slope=1, **Figure 2A**), in agreement with the theoretical estimation of 3 Ct elevation. Surprisingly, the pooling approach did identify many individual samples that had Ct values >37 (**Figure 2C**). A close examination revealed that these cases were typically found in pools that contained ≥2 samples where the viral gene was amplified, and one of the amplified samples had a low Ct (**Figure 2B**). The Ct of a pool is mostly defined by the sample with the highest viral load (lowest Ct) in it; consequently, strongly positive samples lead to individual testing of all samples in the pool, revealing weakly positive “hitchhikers”. The hitchhiker phenomenon explains the better-than-expected sensitivity of our pooling approach. As the average number of positive samples per pool increases, the sensitivity of pooled testing to detect samples with lower viral load (higher Ct) improves (**Figure 2C**). This can be caused by either across the board increase in prevalence, or by clusters of positive samples that are tested in the same pool.

**Figure 2.**
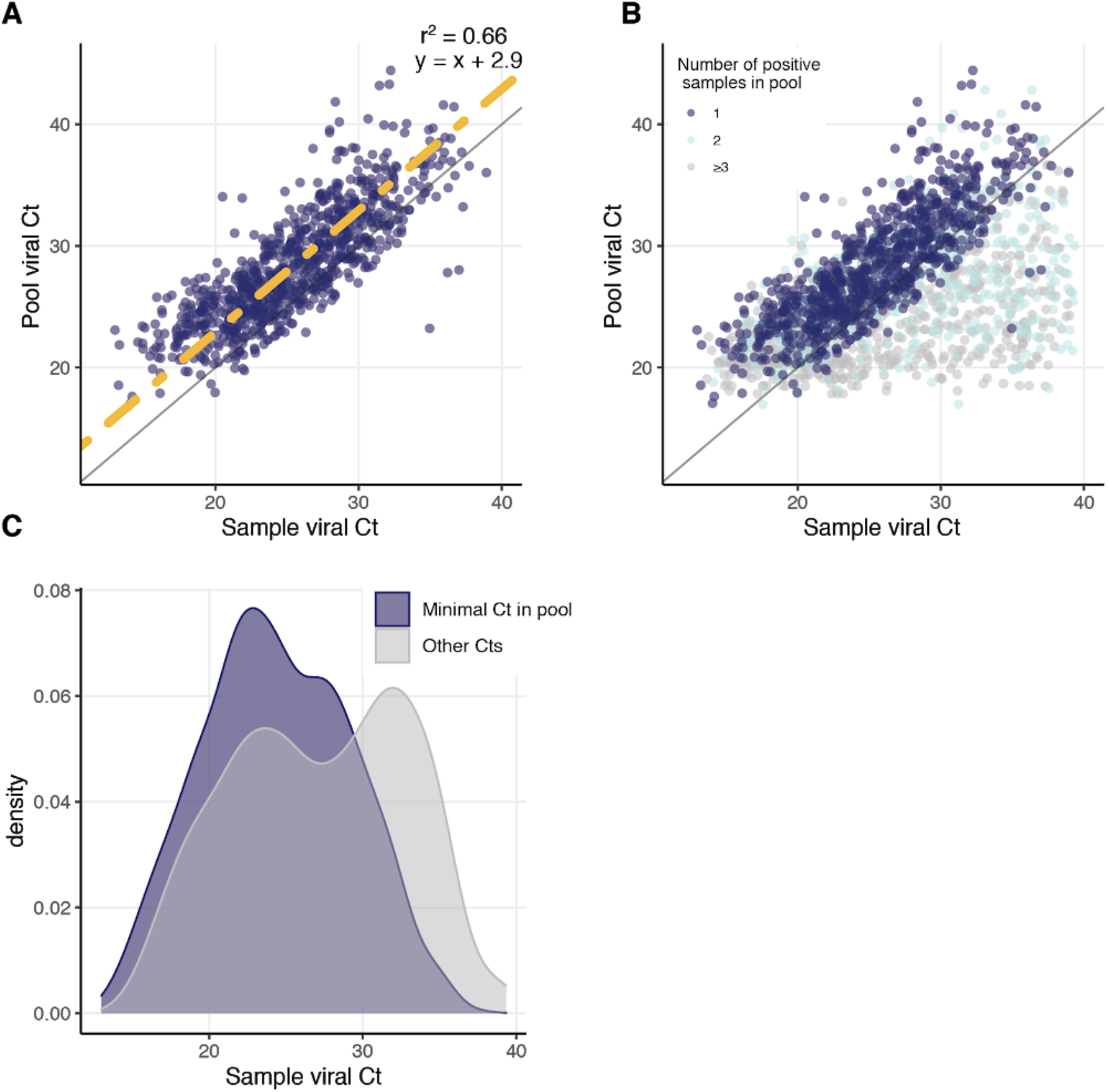
**(A)** Comparison of sample viral Ct (x-axis) and pool viral Ct (y-axis) for all 935 amplified 8-sample pools with a single positive sample. Linear regression with a predetermined slope 1 is marked in yellow, y=x is marked in grey. **(B)** As in (A), including also pools with 241 two amplified samples (light blue) and 82 three or more positive samples (gray). **(C)** Distributions of viral Cts of positive samples in positive pools divided into two groups: sample with the minimal Ct in their pool (blue) and samples with the non-minimal Ct in their pool (gray).

## Discussion

We have employed and monitored a large-scale, adaptive 8- and 5- sample pooling of nasopharyngeal sample lysates for detection of SARS-CoV-2 over a 5-month period. Data analysis of nearly 135,000 pooled samples revealed high empirical efficiency of sample pooling, overweighting a minor clinically-insignificant sensitivity loss. We were able to spare 76% of RNA extraction and RT-PCR tests, even in the reality of frequently changing prevalence rate (<1% to 6%).

Adaptive pooling approaches can maximize resource saving under a fluctuating prevalence rate. The fraction of positive samples tested in pools (*p*) can vary over time (**Figure 1A**) due to multiple factors affecting the epidemic kinetics, including changes in public health mitigation measures (i.e., social distancing regulations, travel restriction, lockdown, school closure)*(19)*. As a result, the pool size (n) required to achieve optimal efficiency shifts. For example, the optimal pool size for *p*=0.02 (2%) is *n*=8, but as *p* rises to 0.05 (5%), optimal pool size shrinks to 5 *(1)* (**Supp Figure 1B**). Consequently, for improved efficiency, we have adopted a dynamic strategy, alternating between pool sizes of 8 and 5, based on the positive rate observed in the past few days, as well as on epidemiological information on the source of samples (e.g. switching to pools of 5 when receiving samples from a source highly suspected to have higher probability of infection) (**Figure 1A, Methods**). Strikingly, we observed supra-optimal empirical efficiency of pooling, exceeding the predicted efficiency, which could not be explained only by the dynamic switching in pool sizes (see below).

When considering the clinical implementation of group testing, loss of sensitivity is a major concern. The dilution of samples due to pooling may lead to lack-of-detection in samples with low viral presence (manifested by high Ct in individual testing). Our empirical results show a loss of sensitivity as expected based on sample dilution. Given the high sensitivity of current SARS-CoV-2 RT-PCR assays, and the accumulated information on the negligible risk of infectiousness associated with low viral RNA level (high Ct), we believe that the loss of 3 Cts is a minor and clinically acceptable tradeoff, as recently suggested*(20)*. Interestingly, our pooling scheme did uncover many positive samples with high Ct values (>37) that would be expected to be missed in pools, presenting real-life performance that exceeds theoretical expectations, similarly to the observed efficiency trend.

We propose that the better-than-expected performance of pooling in both efficiency and sensitivity aspects is rooted in a single factor: the non-random distribution of positive samples in pools. In theory, increased prevalence rates result in decreased efficiency since a common assumption in most models is that samples arrive at random to the diagnostic lab. In reality, samples arrive in batches: from colleges, nursing homes, or healthcare personnel. We sort samples into pools as they arrive at the lab, such that family members and roommates are often pooled together aiming to increase the number of positive samples *within* the pool. The presence of multiple positive samples in a single pool can explain both improved efficiency and improved sensitivity. The efficiency improvement is straightforward: a decision to open a positive pool for individual re-testing results in the discovery of multiple positive samples with the same number of PCR reactions. The sensitivity improvement is less obvious and stems from the relationship between the sample viral Ct and the pool viral Ct (**Figure 2**). A single strongly positive sample is sufficient to make the viral load in the pool detected. If the same pool contains additional low-viral load samples that would have been otherwise missed upon dilution, these would now “benefit” from the higher-viral-load samples co-existing in the pool, and be discovered when the pool is opened for individual testing. Thus, a non-random pool assignment, as well as increased prevalence rate (which by itself increases the likelihood of having pools with multi-positive samples), contribute to the increased sensitivity. A non-random pool assignment together with an adaptive pool size approach further explain our better-than-expected efficiency.

One practical implication of our findings is the importance of using pre-existing knowledge about incoming samples. Using such information for clever co-assignment of samples suspected to be positive or negative can exceed the theoretical performance of pooling typically calculated under the assumption of random assignment. We have encountered considerable logistic hurdles in obtaining pre-test probability for each swab sample, but argue that success in such efforts could make pooling work extremely efficiently even in settings of very high prevalence.

Finally, a common concern with regard to pooling refers to the ease and simplicity of implementation. While some methods may be theoretically more efficient, they need to be manageable at large-scale in a diagnostic lab. We have developed a pipeline that consists of guidelines of which samples to pool, hardware to pool the samples (liquid handlers) and software to pool and track the samples for the second stage of examining individual samples within a positive pool. Details regarding this process including a video demonstrating the entire process can be found in **Supplementary Note 1**.

The long-term containment of Covid-19 will likely involve early identification of outbreaks on the background of very low prevalence in the population. Our empirical evidence from testing over 130,000 samples in pools strongly projects on the feasibility and benefits of carefully-conducted pooling for surveillance, control, and community re-openings.

## Methods

### Sample collection

Nasopharyngeal swab samples were collected in 2 ml Viral Transport Medium (VTM) or directly in the lysis buffer. To inactivate the virus, 220 µL of sample VTM were added to 280 µL 2x Zymo lysis buffer followed by 20′ incubation. For 1:8 pool design we pooled equal volumes of 8 sample lysates to a final volume of 400 µL.

### RNA extraction

RNA was extracted using QIAsymphony DSP Virus/Pathogen Mini kit (Qiagen, Germantown, MD) on Qiasymphony platform and eluted in 60 µL.

### Real-time reverse-transcription PCR

SARS-CoV2 RNA was detected using a multiplex Real-time RT-PCR for the simultaneous detection of SARS-CoV2-specific E gene and a human ERV3 gene as an internal control*(21, 22)*. Primers and probes were purchased from Integrated DNA Technologies (Coralville, IA, USA) and the sequences are described below. Real-time RT-PCR was performed using the TaqPath qPCR Master Mix on the QuantStudio 5 Real-Time PCR Instrument (Applied Biosystems Inc., Foster City, CA).

### Primers and probes used in multiplex Real time RT-PCR

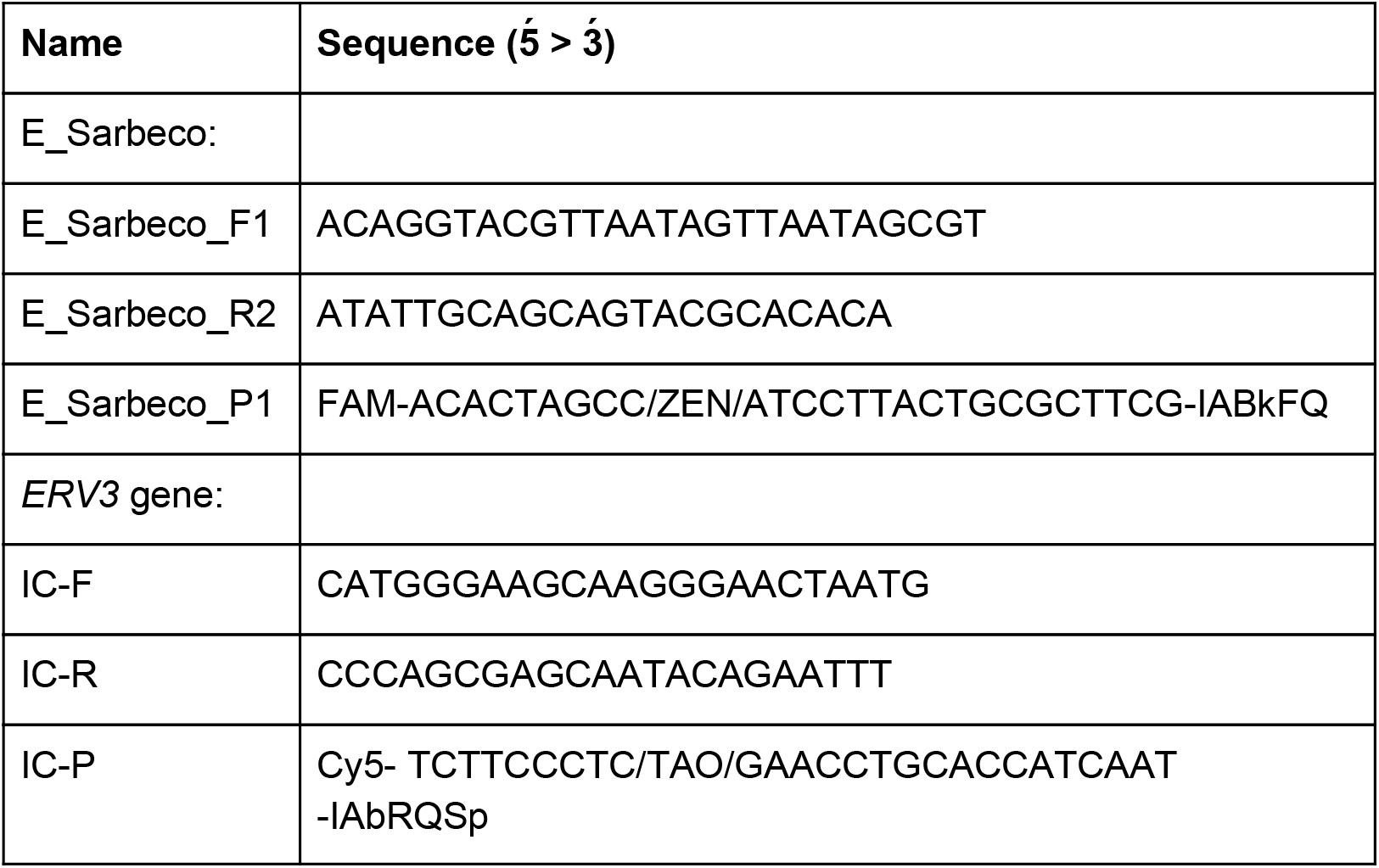

### Definition of positive pools

We based our analysis only on pools that showed amplification of the human gene, used as an internal control. A pool was considered positive if the viral gene was amplified, and individual samples within the pool were re-tested individually.

### Selection of samples for pooling

By and large, samples from symptomatic and hospitalized patients were tested individually, while samples from screened asymptomatic individuals, such as routinely tested hospital personnel and nursing homes residents and caregivers, were pooled.

### Pooling efficiency

When considering Dorfman pooling, for any given assignment of *p* (prevalence rate) and *n* (pool size), the expected dorfman optimal efficiency is calculated as 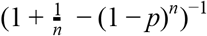, assuming samples are independent and identically distributed across pools*(1)*.

### Pool Ct vs. sample Ct calculation

PCR reaction roughly multiplies the amount of the targeted DNA in each cycle of operation. Due to this exponential growth, a pool of size *n* with a single positive sample should have a Ct that is log2(n) cycles greater than the positive sample’s Ct. For example, when the pool size is 8, this will result in a 3-cycles addition. In order to empirically evaluate the difference between the pool Ct and the single positive sample’s Ct, we applied Linear Regression with a predetermined slope of 1 (**Figure 2A**).

### Practical considerations

We have developed a pipeline that consists of guidelines of which samples to pool, hardware to pool the samples (liquid handlers) and software to pool and track the samples for the second stage of examining individual samples within a positive pool. All details regarding this process including a video demonstrating the entire process can be found in **Supplementary Note 1**.

## Data Availability

Ct values from the pools and individual are available upon request

## Acknowledgements

This research was supported by The Edmond de Rothschild Foundation (Israel) grant for coronavirus research. N. Barak is supported by the Hebrew University Faculty of Medicine Computational Medicine Fellowship. Y. Dor is supported by a generous gift from Shlomo Kramer. M. Yassour is supported by the Azrieli Faculty Fellowship. The following people are members of the Hebrew University-Hadassah Medical School COVID-19 diagnosis team and have contributed to the development of the pooled testing pipeline: A. Klochendler, A. Eden, A. Klar, A. Geldman, A. Arbel, A. Peretz, B. Shalom, B.L. Ochana, D. Avrahami-Tzfati, D. Neiman, D. Steinberg, D. Ben Zvi, E. Shpigel, G. Atlan, H. Klein, H. Chekroun, H. Shani, I. Hazan, I. Ansari, I. Magenheim, J. Moss, J. Magenheim, L. Peretz, L. Feigin, M. Saraby, M. Sherman, M. Bentata, M. Avital, M. Kott, M. Peyser, M. Weitz, M. Shacham, M. Grunewald, N. Sasson, N. Wallis, N. Azazmeh, N. Tzarum, O. Fridlich, R. Sher, R Condiotti, R. Refaeli, R. Ben Ami, R. Zaken-Gallili, R. Helman, S. Ofek, S. Tzaban, S. Piyanzin, S. Anzi, S. Dagan, S. Lilenthal, T. Sido, T. Licht, T. Friehmann, Y. Kaufman, A. Pery, A. Saada, A. Dekel, A. Yeffet, A. Shaag, A. Michael-Gayego, E. Shay, E. Arbib, H. Onallah, K. Ben-Meir, L. Levinzon, L. Cohen-Daniel, L. Natan, M. Hamdan, M. Rivkin, M. Shwieki, O. Vorontsov, R. Barsuk, R. Abramovitch, R. Gutorov, S. Sirhan, S. Abdeen, Y. Yachnin.

---

**Supp Figure 1.**
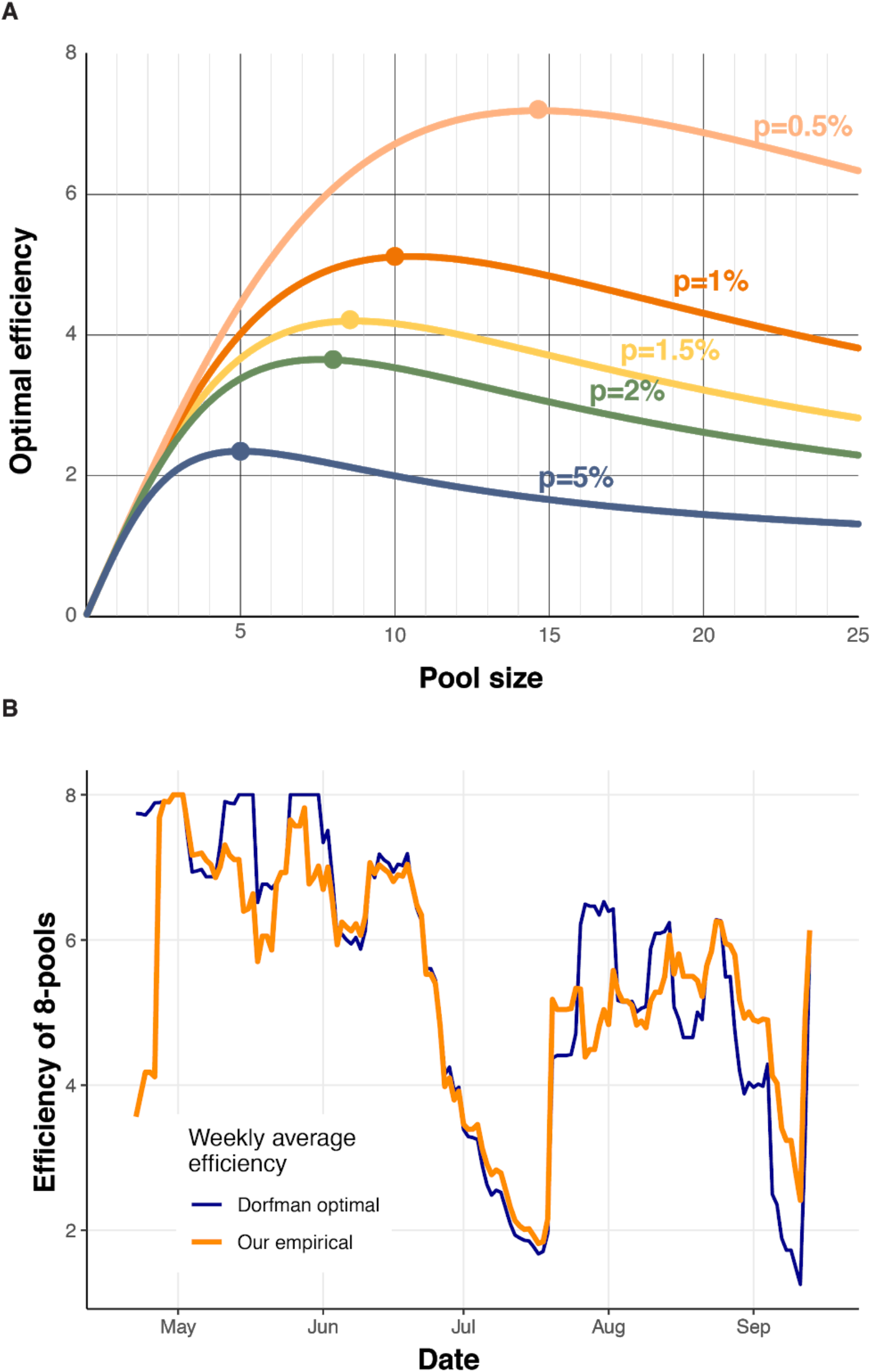
**(A)** Optimal Dorfman efficiency as a function of pool size (x-axis) for varying prevalence rates (colors). **(B)** Weekly average of expected Dorfman efficiency (blue) and our empirical efficiency (orange) for 8-sample pools.

## Supplementary Note 1 – Implementing Dorfman pooling in clinical setting

Successful implementation of high-scale adaptive pooling for SARS-CoV-2 tests requires an appropriate IT infrastructure and automation of information flow. In this supplementary note we will describe the protocol and tools designed and used by the Hadassah Hebrew University COVID-19 diagnosis team. We will address the main challenges and the solutions we used to overcome them.

Unlike individual testing working schemes, pooling requires the ability to **efficiently trace all the individual samples associated with a pool**. We use a *hash file*, created automatically by the liquid handler (LiHa) robot. As a batch of 64 individual samples is pooled into 8 pools, This file links the 8 barcoded individual samples to the corresponding pool barcode. In addition, the date, elution plate barcode, batch number, etc. are automatically added to the file, allowing to quickly locate the individual samples from storage.

Another major challenge is the need **to follow a sample from the time it arrives at the lab and until a test result is reported**. Hence, Hadassah Medical Center IT team adapted the Laboratory Information System (LIS) to support pooling and allow **dynamic pool size selection**. The *hash file and* the results of the PCR test are integrated into the LIS, automatically reporting negative results for all the samples in a negative pool, and assigning all the samples in a positive pool to be retested individually. In addition, lab technicians have a wide set of tools enabling efficient and rapid turnaround such as alerts, data analysis tools for the different stages of pooling, and the ability to compare pooling efficiency for different sample sources.

Our standard operating procedure (SOP) steps are stated and illustrated below, and a video demonstrating the complete pooling procedure can be found here.

1. Prepare *N* individual samples (IS) barcoded tubes containing 500 ul mixture of an individual subject VTM (Viral Transfer Medium) + lysis buffer in each tube.
2. Prepare N/8 empty tubes with a different set of barcodes. These will later contain the pooled samples (PS).
3. Open and load the IS and the empty tubes to a Liquid Handling (LiHa) robot (we use Tecan Freedom Evo 100). Execute Pool Protocol: first 8 IS will be pooled to the 1^st^ PS, the next 8 IS will be pooled to the 2^nd^ PS, etc (50 ul from each, to a total of 400 ul). Alternative faster protocols are available, depending on specifications of the LiHa robot and number of IS.
4. Unload the IS (now containing 450 ul each), close them with new screw caps, and place them in a tube rack, while maintaining their original order on the LiHa robot’s rack. Store them in a safe and marked box (room temperature/4 degrees) until PS PCR results are reported.
5. Check that the *hash file* was created properly and verify each PS is associated with the correct 8 IS barcodes.
6. Unload the PS (now containing 400 ul each), close with new screw caps and transfer to RNA extraction.
7. Perform RNA extraction and RT-PCR on the PS.
8. If the viral Gene in the PS is amplified properly (the PS has viral Ct), locate the relevant IS and validate their barcodes using the *hash file*.
9. Perform RNA extraction and RT-PCR on the suspected IS tubes.

### Process illustration

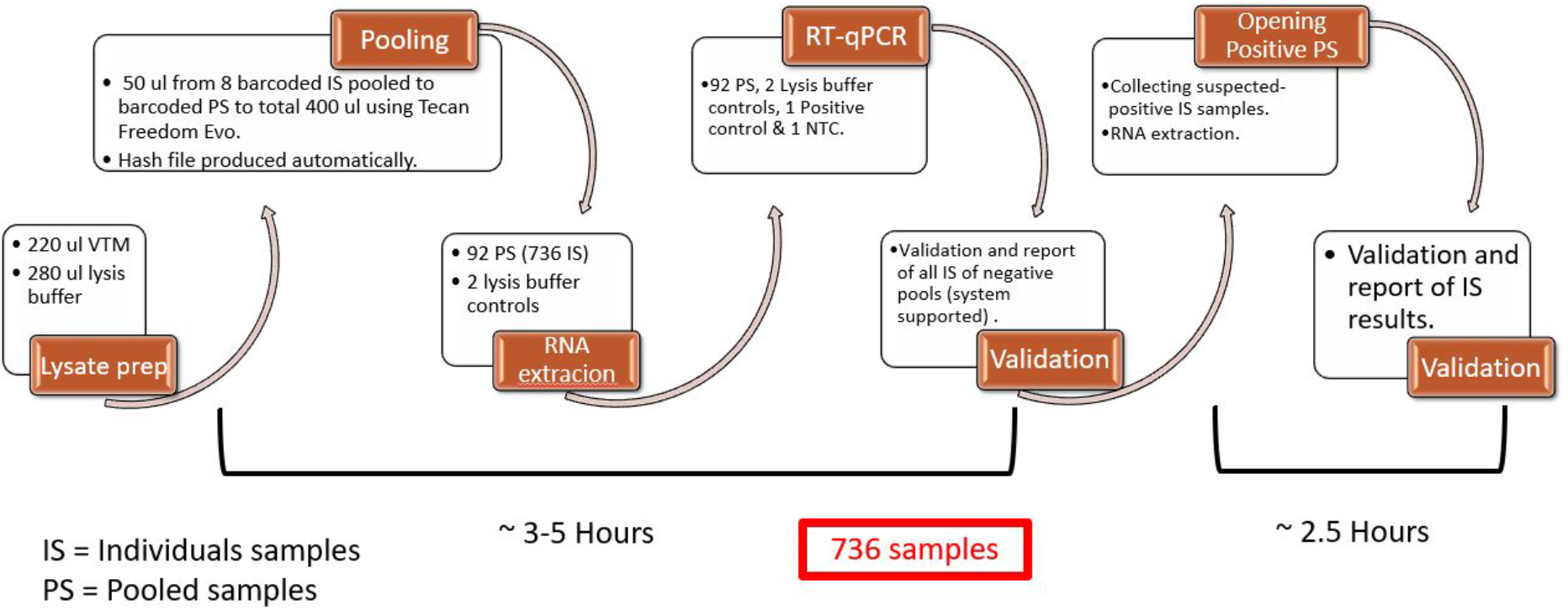

## Notes

### Competing Interest Statement

The authors have declared no competing interest.

### Funding Statement

This research was supported by The Edmond de Rothschild Foundation (Israel) grant for coronavirus research. Y. Dor is supported by a generous gift from Shlomo Kramer. N. Barak is supported by the Hebrew University Faculty of Medicine Computational Medicine Fellowship. M. Yassour is supported by the Azrieli Faculty Fellowship.

### Author Declarations

The study has received IRB exemption from the Israeli Ministry of Health.

